# Modeling rare coding variation on chromosome X provides insight into the genetics and differential sex prevalence of autism spectrum disorder

**DOI:** 10.64898/2026.05.04.26352380

**Authors:** F. Kyle Satterstrom, Kiana Jodeiry, Behrang Mahjani, Gad Hatem, Se Jun Park, Lambertus Klei, Jack M. Fu, Emilie M. Wigdor, the Autism Sequencing Consortium, Catalina Betancur, Mark J. Daly, Kathryn Roeder, Bernie Devlin, Joseph D. Buxbaum, David J. Cutler

**Affiliations:** Stanley Center for Psychiatric Research, Broad Institute of MIT and Harvard, Cambridge, Massachusetts, 02142, USA; Program in Medical and Population Genetics, Broad Institute of MIT and Harvard, Cambridge, Massachusetts, 02142, USA; Analytic and Translational Genetics Unit, Department of Medicine, Massachusetts General Hospital, Boston, Massachusetts, 02114, USA; Department of Psychology, Emory University, Atlanta, Georgia, 30322, USA; Center of Computational and Quantitative Genetics, Emory University School of Medicine, Atlanta, Georgia, 30322, USA; Seaver Autism Center for Research and Treatment, Icahn School of Medicine at Mount Sinai, New York, New York, 10029, USA; Department of Psychiatry, Icahn School of Medicine at Mount Sinai, New York, New York, 10029, USA; Department of Human Genetics, Emory University School of Medicine, Atlanta, Georgia, 30322, USA; Department of Psychiatry, University of Pittsburgh School of Medicine, Pittsburgh, 15213, PA, USA; Center for Genomic Medicine, Massachusetts General Hospital, Boston, 02114, MA, USA; Department of Neurology, Massachusetts General Hospital and Harvard Medical School, Boston, 02114, MA, USA; Department of Paediatrics, University of Oxford, Oxford, OX3 9DU, UK; Institute of Developmental and Regenerative Medicine, University of Oxford, Oxford, OX3 7TY, UK; Institut de Biologie Paris Seine, Sorbonne Université, INSERM, CNRS, Paris, France; Department of Statistics and Data Science, Carnegie Mellon University, Pittsburgh, 15213, PA, USA; Computational Biology Department, Carnegie Mellon University, Pittsburgh, 15213, PA, USA

**Keywords:** Autism Spectrum Disorder, X Chromosome, Sex-Specific Prevalence, Rare Variants

## Abstract

Autism spectrum disorder (ASD) is estimated to be up to four times as common in males as in females, yet the causes of this prevalence difference are not well established. One possible driver is genetic variation on the X chromosome, as it contains genes capable of contributing to ASD (*e.g*., *PTCHD1, MECP2*) and is known to play a role in genetic disorders with differential sex prevalence (*e.g*., color blindness). However, a lack of power compared to the autosomes combined with the complexities of modeling its biology have led to the X being largely overlooked in sequencing studies. Here, we develop quantitative X-linked TADA, a new model designed specifically for application to this chromosome, and use it to analyze rare variation from 50,663 individuals with ASD (and 136,670 individuals total). We find 9 genes on the X associated with ASD at a false discovery rate (FDR) **<** 0.05 and an additional 9 genes at FDR **<** 0.2, with many of these previously identified as involved in specific neurodevelopmental disorders. Point estimates of the liability conferred by *de novo* variants on the X are similar in females and males, with both sexes’ estimates elevated **>**20% above the corresponding autosomal values. We also develop a general theory of how X-linked variation of any additive or non-additive effect influences liability and describe its implications for prevalence. Using this theory and our empirical results, we show how genetic variation on the X could contribute to the sex-differential prevalence of ASD.

## 1 Introduction

Autism spectrum disorder (ASD) is more commonly diagnosed in males than females, with estimates of differential diagnosis near 3:1 or 4:1 [1–4]. The underlying cause of this difference has been investigated, with potential explanations including both biological and social effects [5–13]. For many monogenic disorders with large prevalence differences (*e.g*., color blindness [14]), it is understood that genes on the X chromosome are the primary cause of the difference. However, for complex disorders like ASD, the X has been largely overlooked as a potentially significant factor [15–18]. Although genes on the X are known to contribute to a range of neurodevelopmental disorders [19, 20], the many genes that have been associated with ASD by large-scale sequencing—such as the 72 identified at a false discovery rate (FDR) < 0.001 in our own recent study [21]—are almost exclusively autosomal. We believe that there are at least two reasons that sequencing studies of ASD have not identified more associations on the X.

First, statistical power to detect associations with genes on the X is substantially lower than on the autosomes. This is because rare variant association studies of ASD are generally powered by an observed excess of *de novo* mutations [21, 22], and approximately 80% of all *de novo* mutations occur in the gamete inherited from the father [23]. Females, who inherit an X chromosome from their father, therefore provide much greater power for studies of the X than do males, who inherit a Y chromosome. Yet, because ASD is more common in males than females, ASD studies typically have far more male probands. Thus, a typical ASD study is likely to observe perhaps a third as many *de novo* mutations per gene on the X as on the autosomes, meaning that we would not expect to find ASD associations on the X until study sizes approach three times the size for which autosomal associations were first convincingly demonstrated.

Second, because the X chromosome is hemizygous in males, the role of genetic dominance in disease etiology is fundamentally different on the X than on the autosomes. Most of the analysis tools that have been so successful at discovering autosomal loci (*e.g*., TADA [24], logistic regression) make an assumption of additivity: that the effect of a heterozygous genotype is halfway between the effects of the two homozygotes. While this is a reasonable assumption when studying rare (*q* ≤ 10^−3^) variation, as such variation is largely unobserved in autosomal homozygous form (*q*^2^ ≤ 10^−6^) at current study sizes, it means that when alleles are observed in unaffected individuals their presence is taken as evidence against association. On the X, by contrast, a rare highly penetrant recessive allele might never be observed as a homozygous genotype in any female, but such a variant will be observed hemizygously in affected males and heterozygously in *unaffected* females.

The biology of the X chromosome thus requires adjustment of existing methods. To account for these differences, we have developed a new tool, quantitative X-linked TADA (QXL-TADA), which differs from our previous TADA model [21, 22, 24] by utililzing the liability scale and including multiple disease models to account for possibilities introduced by the biology of the X. This model can be used to study the genetics of the X chromosome in any phenotype. Here, we apply it to rare coding variants from a large cohort of individuals with ASD, and we demonstrate substantial evidence for the association of several genes on the X. We suggest that many more genes on the X may contribute to ASD as well. Finally, we describe the quantitative genetics theory by which variants in these genes could contribute to the observed ASD prevalence difference between the sexes.

## 2 Results

### 2.1 ASD cohort and *de novo* variant rates

We analyze rare variants from 50,663 individuals with ASD and 36,498 individuals without ASD (Table 1). These numbers include 26,738 ASD probands (21,332 male and 5,406 female) and 9,567 unaffected siblings (4,739 male and 4,828 female) for whom we have parental sequences and can thus identify *de novo* variants, with the remaining samples being unrelated cases and controls. Autosomal variants from approximately 40% of these samples were analyzed in our previously published work [21]. We classify variants according to their predicted effect: putative protein-truncating variants (PTVs, including stop gained, frameshift, and splice donor/acceptor variants); missense variants predicted to have a deleterious impact (MisB), missense variants predicted to have a moderate impact (MisA); all missense variants (All Mis), regardless of predicted effect; and synonymous variants (Syn). The missense classes are based on the value of a variant’s “Missense badness, PolyPhen-2, and Constraint” (MPC) estimator of deleteriousness [25], where increasing scores can be thought of as increasing confidence that the variant has an effect on protein function. Following our previous work [21, 22], we take MisB to be MPC ≥ 2 and MisA to be 2 > MPC ≥ 1.

**Table 1:** Sample counts by data type and diagnosis category. Probands and cases are individuals with recorded ASD, and siblings and controls are those without. Probands and siblings have also had parents sequenced, enabling the distinction between *de novo* and inherited variants.

|  | Family-Based |  |  |  | Case-Control |  |  |  |
| --- | --- | --- | --- | --- | --- | --- | --- | --- |
|  | Proband |  | Sibling |  | Case |  | Control |  |
|  | Male | Female | Male | Female | Male | Female | Male | Female |
|  | 21,332 | 5,406 | 4,739 | 4,828 | 16,841 | 7,084 | 17,816 | 9,115 |
| Totals | 26,738 |  | 9,567 |  | 23,925 |  | 26,931 |  |

Consistent with our expectations for X-linked biology, we observe vastly more *de novo* variants on the X per female (probands and siblings combined) than per male, regardless of variant class (Table 2; our analysis does not include the pseudoautosomal regions (PAR)). For example, for synonymous sites, we observe 110 *de novo* variants in females and 46 in males, a female/male ratio of approximately 6.1 when adjusted for sample size (or 3.0 when adjusted for copy number). The female to male *de novo* excess is similar for the other classes of mutation considered, with each female contributing between 6.0 and 7.0 times as many variants as each male.

**Table 2:** *De novo* variant rates by sample group. Values outside parentheses are for the X chromosome (and values inside are for the autosomes). *De Novo* is the total number of events observed, and Rate is the rate per gene, adjusted for sex-specific copy number. GRR is the genotype relative risk, defined by the ratio of the rate in affected to unaffected individuals, and P is the Fisher’s exact test p-value comparing counts in ASD probands to unaffected siblings. Calculations assume 816 chromosome X genes and 19,478 autosomal genes. Results do not include the pseudoautosomal regions of the X.

| Variant Class | Sex | Affectation | *De Novo* X (Auto) | Rate X (Auto) | GRR X (Auto) | P X (Auto) |
| --- | --- | --- | --- | --- | --- | --- |
| PTV | Male | Proband | 28 (2920) | $1.61 \times 10^{-6}$ ($3.51 \times 10^{-6}$) | 1.04 (1.44) | 1 ($3.88 \times 10^{-14}$) |
| | Male | Sibling | 6 (449) | $1.55 \times 10^{-6}$ ($2.43 \times 10^{-6}$) | | |
| | Female | Proband | 57 (878) | $6.46 \times 10^{-6}$ ($4.17 \times 10^{-6}$) | 2.04 (1.77) | $2.64 \times 10^{-3}$ ($1.04 \times 10^{-23}$) |
| | Female | Sibling | 25 (443) | $3.17 \times 10^{-6}$ ($2.36 \times 10^{-6}$) | | |
| MisB | Male | Proband | 23 (1169) | $1.32 \times 10^{-6}$ ($1.41 \times 10^{-6}$) | 2.55 (1.55) | 0.30 ($3.55 \times 10^{-8}$) |
| | Male | Sibling | 2 (168) | $5.17 \times 10^{-7}$ ($9.10 \times 10^{-7}$) | | |
| | Female | Proband | 48 (342) | $5.44 \times 10^{-6}$ ($1.62 \times 10^{-6}$) | 2.86 (1.86) | $1.99 \times 10^{-4}$ ($1.76 \times 10^{-11}$) |
| | Female | Sibling | 15 (164) | $1.90 \times 10^{-6}$ ($8.72 \times 10^{-7}$) | | |
| MisA | Male | Proband | 28 (3022) | $1.61 \times 10^{-6}$ ($3.64 \times 10^{-6}$) | 1.24 (1.18) | 0.82 ($2.54 \times 10^{-4}$) |
| | Male | Sibling | 5 (569) | $1.29 \times 10^{-6}$ ($3.08 \times 10^{-6}$) | | |
| | Female | Proband | 47 (770) | $5.33 \times 10^{-6}$ ($3.66 \times 10^{-6}$) | 1.35 (1.05) | 0.21 (0.41) |
| | Female | Sibling | 31 (658) | $3.93 \times 10^{-6}$ ($3.50 \times 10^{-6}$) | | |
| All Mis | Male | Proband | 105 (15955) | $6.03 \times 10^{-6}$ ($1.92 \times 10^{-5}$) | 1.30 (1.08) | 0.35 ($8.00 \times 10^{-5}$) |
| | Male | Sibling | 18 (3288) | $4.65 \times 10^{-6}$ ($1.78 \times 10^{-5}$) | | |
| | Female | Proband | 219 (4107) | $2.48 \times 10^{-5}$ ($1.95 \times 10^{-5}$) | 1.62 (1.05) | $1.67 \times 10^{-5}$ (0.02) |
| | Female | Sibling | 121 (3477) | $1.54 \times 10^{-5}$ ($1.85 \times 10^{-5}$) | | |
| Syn | Male | Proband | 40 (6191) | $2.30 \times 10^{-6}$ ($7.45 \times 10^{-6}$) | 1.48 (1.02) | 0.45 (0.61) |
| | Male | Sibling | 6 (1354) | $1.55 \times 10^{-6}$ ($7.33 \times 10^{-6}$) | | |
| | Female | Proband | 64 (1590) | $7.25 \times 10^{-6}$ ($7.55 \times 10^{-6}$) | 1.24 (1.04) | 0.29 (0.25) |
| | Female | Sibling | 46 (1360) | $5.84 \times 10^{-6}$ ($7.23 \times 10^{-6}$) | | |

Comparing the rates in probands and siblings within each sex (calculating the genotype relative risk, or GRR), we find significant enrichment on the X in female probands for *de novo* PTVs, MisB variants, and All Mis variants (Table 2). Although none of the comparisons are significant for males, point estimates of the GRR are similar to females across the missense classes, suggesting that the lack of significance is likely due to observing fewer mutations in males. We note that the GRR for PTVs on the X would increase to 1.28 in males (and 2.74 in females) if restricted to only constrained genes (*i.e*., the bottom three deciles of LOEUF [26]). Although we do observe a higher rate of *de novo* synonymous mutations on the X in male probands as compared to male siblings, this relies—like the other variant classes in males—on very small numbers; we call only six synonymous *de novo* variants across all male siblings, so that the difference in rates is sensitive to even a single additional mutation.

### 2.2 QXL-TADA and the liability scale

QXL-TADA, which we employ for further analysis of our cohort, models the distribution of disease risk and prevalence in a population using the mixed model of inheritance of Morton and MacLean [27] (for details, see Methods, Supplemental Methods, and https://gitlab.com/gadhatem/david_cutler_lab_x_chromosome). In this framework, each individual has some underlying quantitative liability of developing the outcome. If we assume that the genetic and environmental factors contributing to an individual’s liability are independent and additive, then liability is approximately normally distributed in the population. Individuals with liabilities above some threshold are affected, and individuals with liabilities below that threshold are unaffected (Figure 1A). For a biallelic gene, the population liability distribution can be dissected into three curves, representing liability distributions for individuals with each of the three genotypes (Figure 1B). Sex-specific differences in allele penetrance are modeled mathematically as differing thresholds on the liability scale (Figure 1C).

**Fig. 1:**
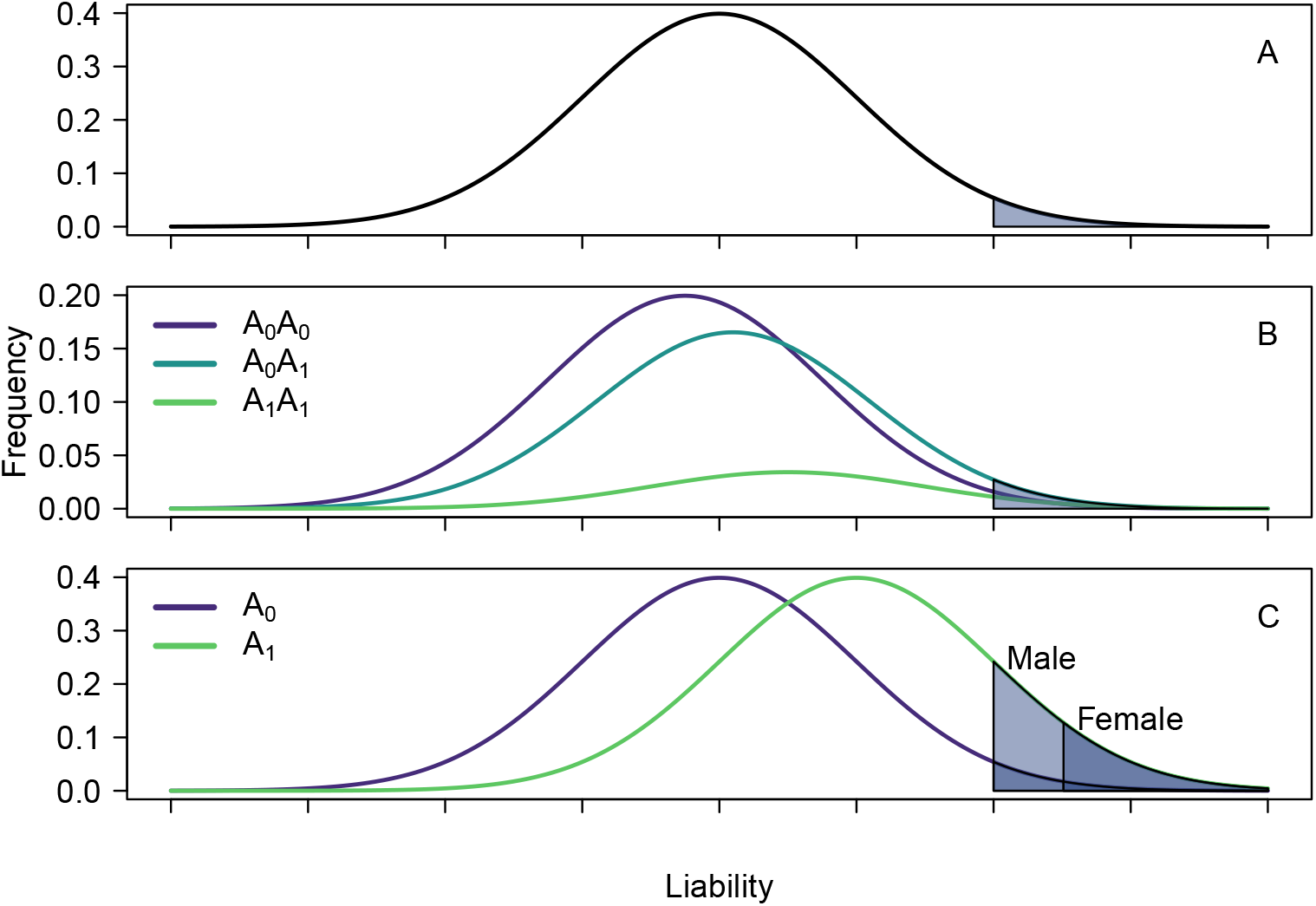
Example liability distributions. A) Binary traits such as ASD are modeled as thresholds on a normally distributed scale called liability. A threshold is shown, above which individuals are affected. B) Genotypes with differing mean liability have differing penetrances, determined as the fraction of the liability distribution beyond the threshold. C) Sex-specific prevalence differences can be modeled as differing thresholds on the liability scale. This necessarily implies differing penetrances of alleles of large effect, such as example allele *A*_1_, and therefore differing odds ratios between the sexes.

Like the original TADA, QXL-TADA is a gene-based test. Within each gene, different variants of the same class are assumed to have the same effect on the liability scale and are modeled as a single allele. Correspondingly, each sample’s genotypes across all variants of a given class within a gene are summarized by a single “gene-level genotype” label. On the X chromosome, males with one or more variants of a class are labeled hemizygotes. Females with one variant are labeled heterozygotes, and females with two or more variants are labeled homozygotes. An allele possessed by a child is counted as *de novo* if it could not have been inherited from a parent (*e.g*., if the child had a PTV in a gene where neither parent had one).

On the liability scale, estimates of effect sizes in males and females are similar across variant classes (Figure 2), with point estimates consistently >20% larger on the X than on the autosomes for both sexes. Although the confidence intervals in the male data are too wide to draw strong conclusions, these estimates suggest that the average mutation on the X could confer more liability than the average autosomal mutation, or that a larger fraction of genes on the X could be involved in risk. Overall, looking across sexes and variant classes, our data support the idea that previous sequencing studies of ASD have found few associations on the X not because effect sizes are lower but rather because power is decreased.

**Fig. 2:**
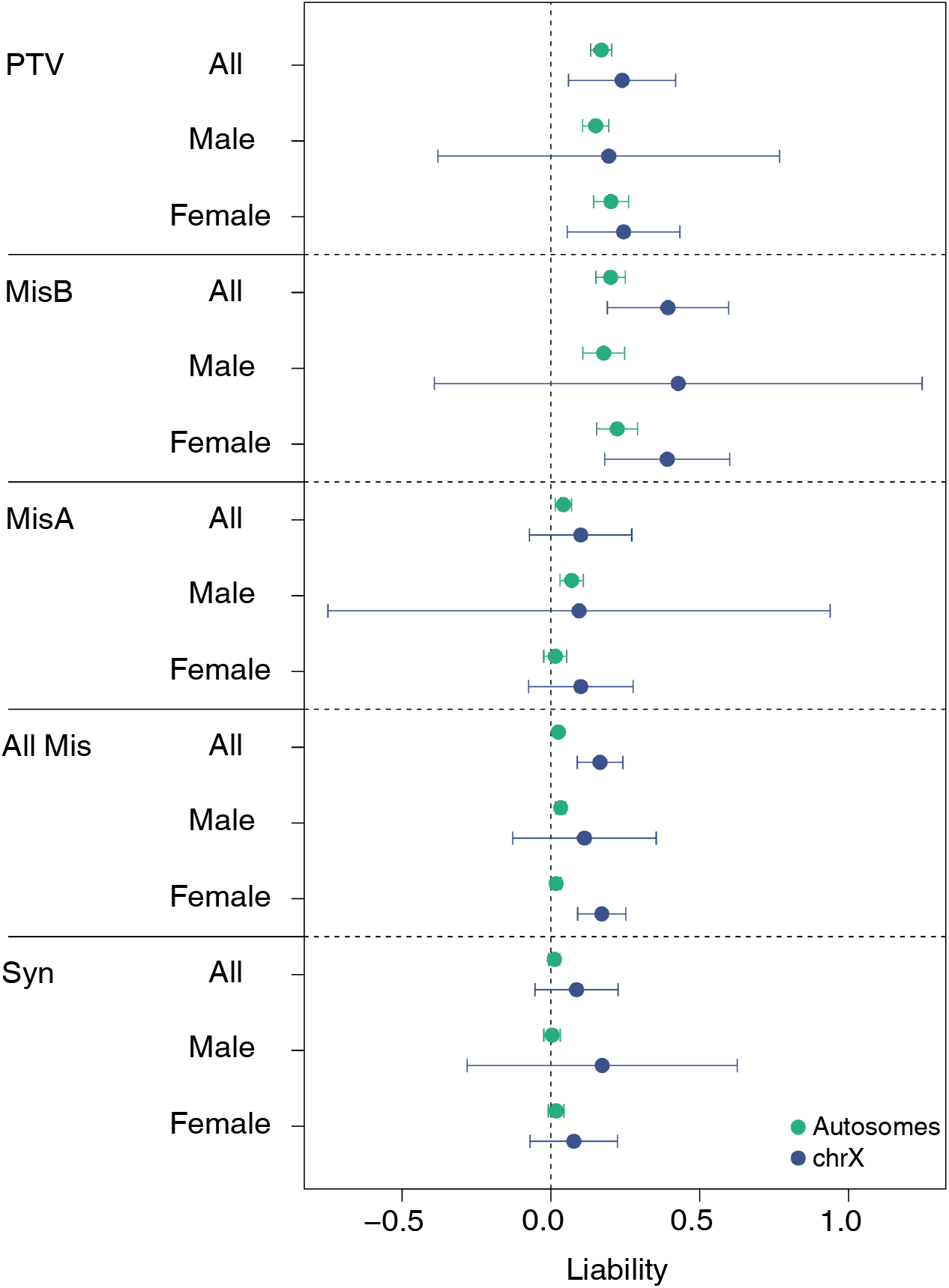
Liability conferred across *de novo* variant classes and sexes in our ASD cohort. Estimates are shown for genes on the autosomes (green) and the X (blue). Due to vastly different sample sizes (there are more autosomal genes, and males contribute more mutations per gene on the autosomes than the X), confidence intervals are much wider for the X than the autosomes. In females, there is statistically significant enrichment for *de novo* PTVs, MisB variants, and All Mis variants on the X. These variant classes continue to have larger point estimates of their average effect on the X than the autosomes when both sexes are combined (PTVs = 0.24 vs 0.17; MisB = 0.40 vs 0.20; All Mis = 0.17 vs 0.025). There is no statistically significant enrichment in males for any class on the X, but point estimates in males and females are largely consistent. Error bars reflect twice the standard error.

### 2.3 Gene-level association results for ASD

We apply QXL-TADA to all of the ASD samples for which we have variants (Table 1), including family-based data and case-control data. For each gene and variant class, all data, regardless of cohort and family status, are analyzed simultaneously. Observed gene-level genotype counts are modeled as independent draws from a multinomial distribution defined by several parameters to account for potential population structure and demographic confounding (Supplemental Methods). For comparison to the Null model (where the variant allele has no effect on the outcome), we consider three disease models: (i) an Additive effect model (female heterozygotes have half the liability of female homozygotes or male hemizygotes), (ii) a Recessive effect model (only male hemizygotes or female homozygotes have any effect), and (iii) a Homozygous Lethal model (female heterozygotes have an effect, but male hemizygotes and female homozygotes are unobserved). These models are compared to the Null via a likelihood ratio test (LRT), and we construct one gene-level p-value by combining the LRT p-values from the three disease models with a truncated Cauchy combination test (CCT) [28]. On simulated data, this method does not show inflation (Figures S2 and S3).

We test each of our five classes of variation for association with ASD, considering each non-PAR chromosome X gene where we observe any variants of the given class. To guard against false positives, we drop 65 genes from the PTV analysis due to >50% of their estimated PTV mutability coming from sites that do not pass loftee [26] filters (Methods). We observe in the resulting quantile-quantile plots that CCT p-values are clearly enriched above the null for PTVs, are somewhat enriched for MisB variants, and show essentially no signal for MisA variants, All Mis variants, or Syn variants (Figure 3). The overall fraction of null loci (estimated using the convest algorithm implemented in the propTrueNull function of the R limma package) is consistent with this; only the PTV (18.2% genes estimated non-null) and MisB (4.5% genes estimated non-null) classes show the potential for a substantial number of genes with rare variation contributing to ASD. Previous studies have estimated that roughly 1,000 autosomal genes could harbor rare alleles that contribute to ASD [22, 24], or ~6% of the autosomal genes queried by exome sequencing; our results suggest that a substantially larger proportion of genes on the X could be involved. Lambda-GC values for our data (0.78 for PTVs; 0.21 for MisB; 0.04 for MisA; 0.01 for All Mis; and 0.001 for Syn) do not suggest overall test statistic inflation.

**Fig. 3:**
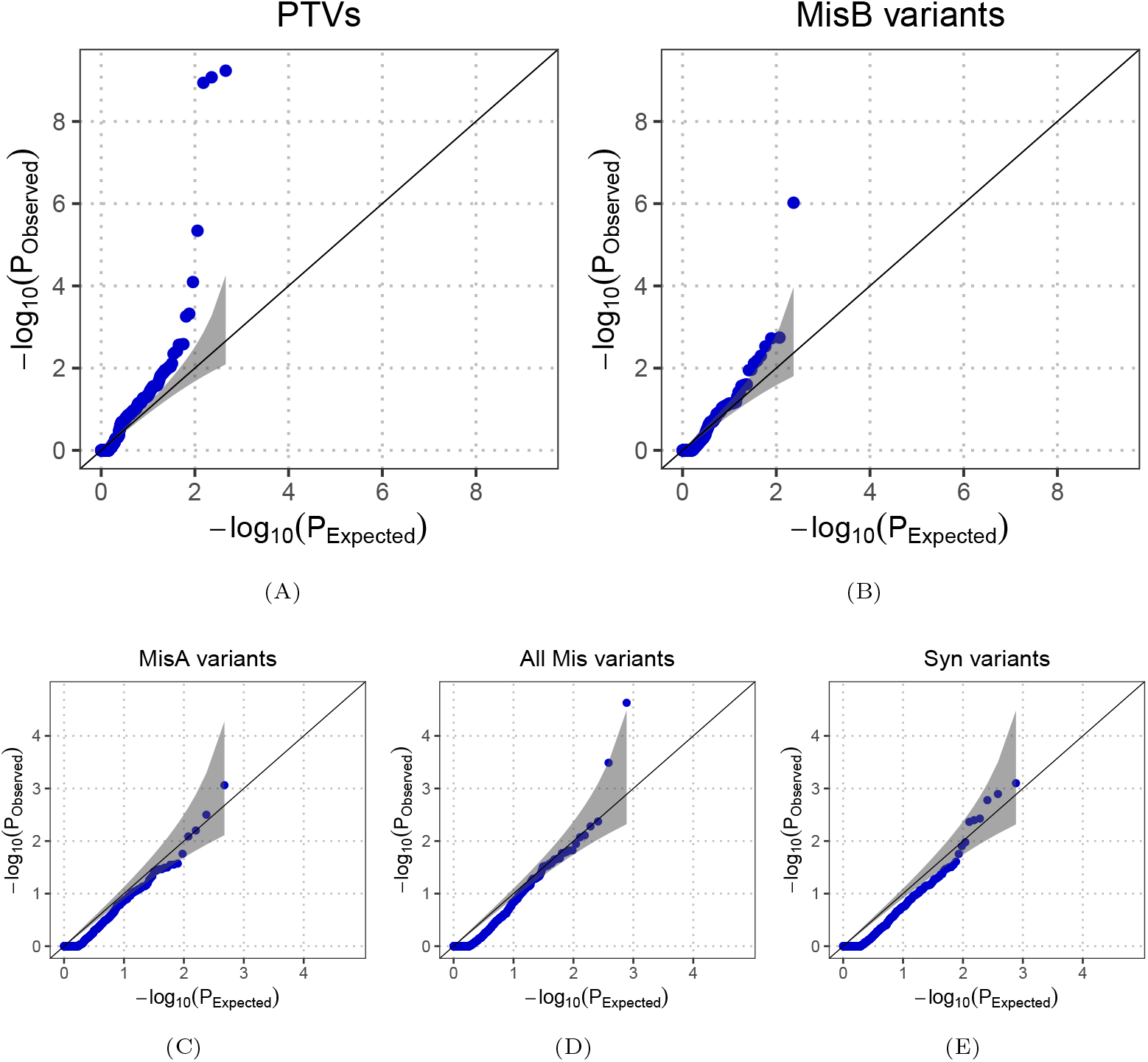
Quantile-quantile plots of QXL-TADA results. Each dot represents the Cauchy combination test p-value for an X-linked gene tested for ASD association via rare alleles of the given class of variation. Only genes with at least one variant of the given type are plotted. The estimated proportions of true null loci are 3A) PTV: 81.8%, 3B) MisB: 95.5%, 3D) MisA: 100%, 3C) All Mis: 100%, and 3E) Synonymous: 99.9%. These estimates are influenced by the excess of genes with *p* ≈ 1.0 (due to genes with only a small number of variants being underpowered to distinguish from the Null model) in all variant classes.

We next calculate Benjamini-Hochberg FDR values for each combination of gene and variant class, noting the best-fitting disease model. We find nine genes with FDR < 0.05 and another nine with FDR < 0.2, with PTVs contributing the majority of the associations and the MisB and All Mis classes contributing the remainder (Table 3). Existing literature provides broad support for our top results, as eight of the nine genes identified by QXL-TADA at FDR < 0.05 have “definitive” or “strong” evidence for association with developmental disorders (DD) in the curated Gene2Phenotype (G2P) database (https://www.ebi.ac.uk/gene2phenotype/panel/DD [29]), while the remaining gene (*TCEAL1*) displays “moderate” evidence. Like *MECP2* and Rett syndrome [30], they have each been identified as underlying specific neurodevelopmental disorders associated with intellectual disability [31–38]. In addition, each gene we identify through PTVs at this FDR threshold is also associated with some level of seizures [32, 34–36, 39–41], providing support for a relationship between autism and epilepsy. Among genes with FDR values between 0.05 and 0.2, *NLGN4X* [42], *WDR45* [43], *FTSJ1* [44], *SYP* [45], and *TFE3* [46] all appear in the G2P DD panel with “definitive” or “strong” support, and there is “limited” evidence for *CDK16* [47]. *IL1RAPL2* is a homolog of *IL1RAPL1*, a known intellectual disability gene [48]. QXL-TADA estimates penetrance values as part of its analysis, and these values are above 95% for the rare allele hemizygous male / homozygous female for 8 of the 13 genes listed in Table 3 when the best-fitting model is Additive or Recessive; estimated penetrance is above 50% for heterozygous females for *DDX3X* and the Homozygous Lethal model. Somewhat reassuringly this is consistent with previous observations of *DDX3X* where PTV variants appear highly penetrant in heterozygous females and absent in homozygous or hemizyous state. Furthermore, we note that MisB variants in *DDX3X* show more than nominal association (*p* = 0.0066), with an additive model fitting best, also consistent with prior observations, although our confidence in this association is below our reporting threshold (FDR = 0.256). We do not identify any genes with FDR < 0.2 for the MisA or Syn variant classes.

**Table 3:** Chromosome X genes associated with ASD at two FDR thresholds. Model column gives the disease model with the greatest evidence.

| Gene | FDR < 0.05 | Model | Gene | 0.05 < FDR < 0.2 | Model |
| --- | --- | --- | --- | --- | --- |
|  | Variant Class |  |  | Variant Class |  |
| *DDX3X* | PTV | Lethal | *NLGN4X* | PTV | Recessive |
| *MECP2* | PTV | Additive | *WDR45* | PTV | Lethal |
| *ARHGEF9* | PTV | Additive | *CDK16* | PTV | Recessive |
| *IQSEC2* | PTV | Additive | *FTSJ1* | PTV | Recessive |
| *CNKSR2* | PTV | Additive | *SLC6A14* | PTV | Lethal |
| *TCEAL1* | PTV | Additive | *SYP* | PTV | Recessive |
| *CLCN4* | PTV | Recessive | *TFE3* | MisB | Lethal |
| *NAA10* | MisB | Additive | *IL1RAPL2* | MisB | Additive |
| *IDS* | All Mis | Recessive | *SSX4* | All Mis | Lethal |

**Table 4:**
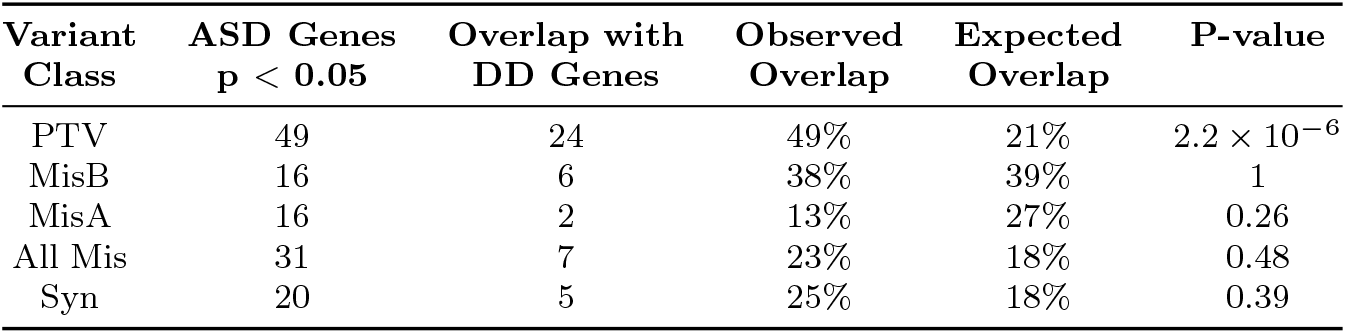
Overlap of ASD genes with developmental disorders (DD) genes on the X. DD genes include 140 non-PAR chromosome X genes with “definitive” or “strong” evidence in the G2P DD panel [29]. Expected values reflect the fraction of genes with variation of a given class in our analysis that overlap the DD genes. P-values are by Fisher’s exact test.

| Variant Class | ASD Genes<br>$p < 0.05$ | Overlap with<br>DD Genes | Observed<br>Overlap | Expected<br>Overlap | P-value |
| --- | --- | --- | --- | --- | --- |
| PTV | 49 | 24 | 49% | 21% | $2.2 \times 10^{-6}$ |
| MisB | 16 | 6 | 38% | 39% | 1 |
| MisA | 16 | 2 | 13% | 27% | 0.26 |
| All Mis | 31 | 7 | 23% | 18% | 0.48 |
| Syn | 20 | 5 | 25% | 18% | 0.39 |

**Table 5:** Sample counts for family-based and case-control data by dataset.

| Cohort | Family-Based |  |  |  | Case-Control |  |  |  |
| --- | --- | --- | --- | --- | --- | --- | --- | --- |
|  | Proband |  | Sibling |  | Case |  | Control |  |
|  | Male | Female | Male | Female | Male | Female | Male | Female |
| ASC B14 | 6,025 | 1,265 | 1,158 | 1,190 |  |  |  |  |
| ASC B15-16 | 223 | 56 | 6 | 5 |  |  |  |  |
| SPARK Pilot | 376 | 89 |  |  |  |  |  |  |
| SPARK WES1 GATK | 5,219 | 1,324 | 1,554 | 1,478 |  |  |  |  |
| iPSYCH |  |  |  |  | 3,730 | 1,133 | 3,373 | 1,629 |
| PAGES |  |  |  |  | 518 | 210 | 1,850 | 1,745 |
| ASC B17-21 | 887 | 222 | 27 | 13 |  |  |  |  |
| SPARK iWES v2 | 8,602 | 2,450 | 1,994 | 2,142 |  |  |  |  |
| SPARK-UKB |  |  |  |  | 12,593 | 5,741 | 12,593 | 5,741 |
| Totals: By Sex | 21,332 | 5,406 | 4,739 | 4,828 | 16,841 | 7,084 | 17,816 | 9,115 |
| Totals: Combined | 26,738 |  | 9,567 |  | 23,925 |  | 26,931 |  |

To better understand the relationship between genes on the X associated with ASD and those involved in the broader spectrum of DD phenotypes, we examined the overlap between genes with at least nominal significance (CCT p-value < 0.05) in our analysis and the 140 non-PAR chromosome X genes with “definitive” or “strong” evidence from the G2P DD panel [29]. Genes identified in our study via PTVs or MisB variants substantially overlapped the DD genes, although without statistical significance for the MisB class (the expectation for MisB overlap is higher, as only genes with missense constrained regions—and therefore some evidence for the impact of mutation on fitness—are capable of variation in this category). For the other variant classes, the overlap between DD and ASD genes is not significantly different than chance. While it could be coincidence, it is striking that the baseline expectation is set by G2P [29] identifying ~18% of genes on the X as being involved in DDs, and our PTV non-null estimate is also ~18%.

### 2.4 Implications for sex-differential prevalence of ASD and other binary phenotypes

QXL-TADA and heritability studies in general model binary traits as resulting from a threshold on an unobserved normally distributed liability scale [49]. On this scale, there are four models that could explain between-sex differences in prevalence: differences in liability threshold (Figure 4A), differences in mean liability (Figure 4B), differences in its variance (Figure 4C), or differences in both mean and variance (Figure 4D). If all variation were autosomal, the first two of these models would be mathematically indistinguishable. Most heritability studies assume liability to have mean 0 and variance 1 for both males and females, and any prevalence difference is assumed to result from different thresholds on the liability scale. Under this model, ASD prevalences of 2.97% for males and 0.69% for females [2] imply thresholds of 1.885 and 2.462, respectively (Figure 4A). However, if there is variation on the X contributing, the distinctions between the liability models can be fundamental to our understanding of how sex-differential prevalences arise. This is because all X loci contribute to variance differences between males and females, and any X locus with non-additive effects (*i.e*., some form of dominance) contributes to mean differences between males and females.

**Fig. 4:**
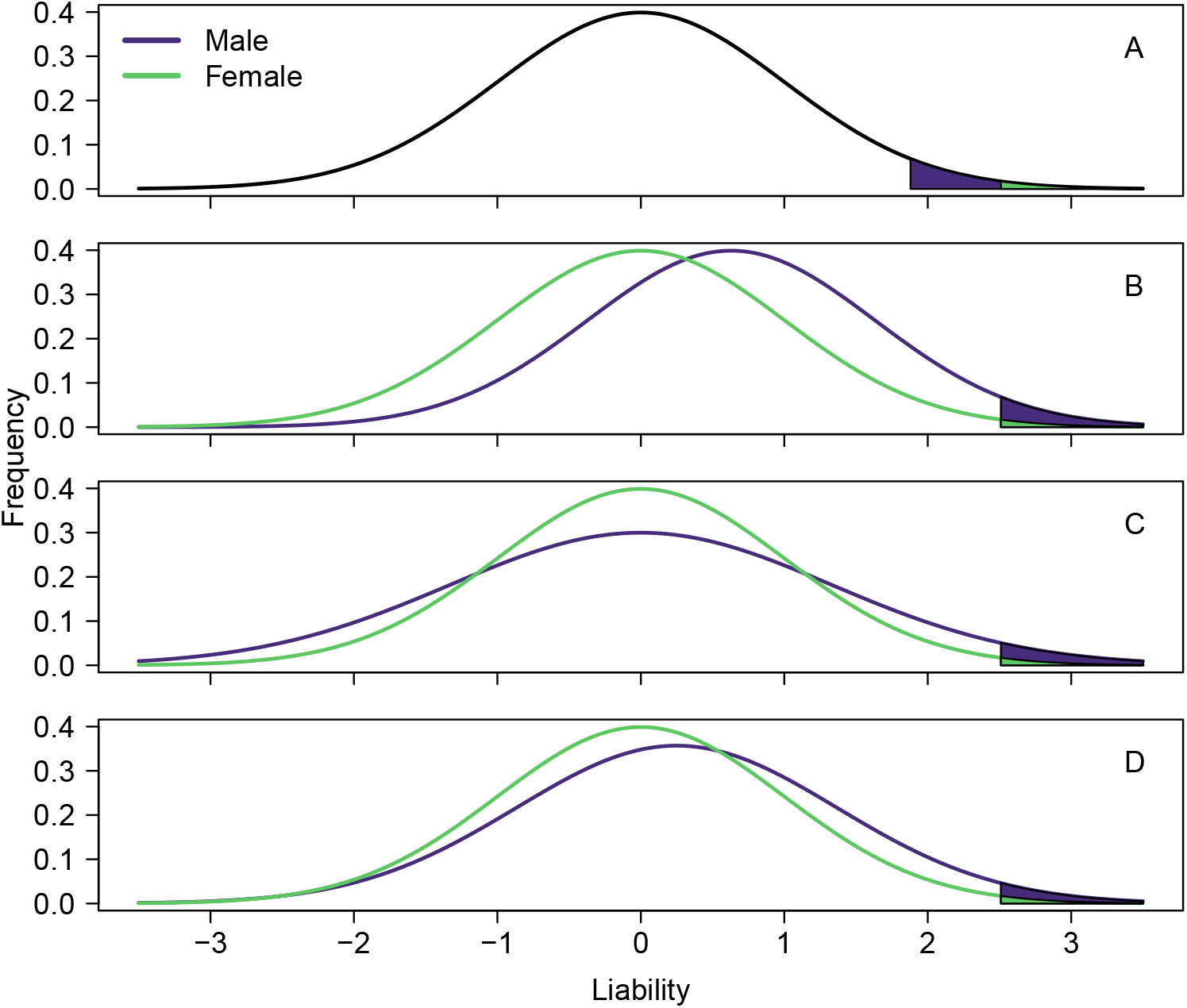
Models for differential male and female prevalence. A) Males and females both have liability mean 0 and variance 1, while males have threshold 1.885 and females have threshold 2.462. B) Males have liability mean 0.577 while females have mean 0. Both have variance 1 and threshold 2.462. C) Males and females both have mean 0 and threshold 2.462, while males have variance 1.71 and females have variance 1. D) Threshold is 2.462 in both males and females. Males have a mean and variance both greater than females.

All variation on the X chromosome contributes to males and females having different amounts genetic variation for a trait. Measured on the liability scale, an X-linked variant contributes ^*f*^ *V*_*g*_ variance to females and ^*m*^*V*_*g*_ to males. (Methods, Supplemental Methods). For a variant on the X with minor allele frequency *q*, major allele frequency *p* = 1 − *q*, additive effect *β*, and dominance deviation *δ* (*i.e*., the deviation of the heterozygote phenotype from additivity), these contributions are:

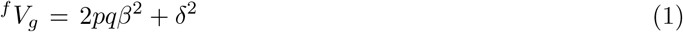

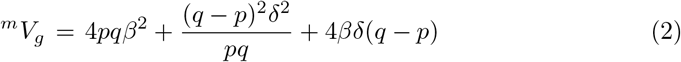

Additionally, any variant that shows any degree of dominance *δ* ≠ 0 contributes −*δ* to the male mean (see Methods for derivation). Thus, while mean liability is always equal to zero in females, it is only equal to zero in males if all X loci are additive. Any locus on the X chromosome where the disease-associated allele is recessive contributes to a positive mean in males, and this locus acts to increase male prevalence of disease relative to female prevalence. Thus, X-linked loci necessarily contribute to variance differences between males and females, as well as mean differences, for all non-additive loci.

Given these theoretical results, could results from our empirical analysis explain the observed sex-specific prevalence of autism? In our study, we focused on rare variants in an effort to enrich for alleles of large effect, and we found that *de novo* PTVs on the X had an average effect on the liabilty scale of ~0.24 (Figure 2). If we were to make the broad assumption that any rare PTV had this effect across all genes on the X, their average frequency over all X-linked genes would need to be ~4 × 10^−3^ (if mostly additive) or ~1 × 10^−3^ (if mostly recessive) per gene/chromosome in order to account for the prevalence difference themselves. These are unrealistically large values, as the overall frequency of rare PTVs observed in this study is closer to 10^−5^ per gene/chromosome. At the observed frequency, PTVs on the X would account for < 1% of the observed male-female prevalence difference, contributing approximately 0.2% of ASD heritability in females and somewhat more than 0.4% in males (depending on the rate of dominance). Thus, rare variants on their own are unlikely to make a large contribution to the observed prevalence difference.

In fact, our framework shows that if genetics were to explain a substantial amount of the prevalence difference, the contributing variants would need to be much more common than the ones in this study. This is perhaps not impossible, considering that (1) in this work we estimate a non-null PTV fraction of 18.2%, suggesting that as many as 150 X-linked genes could contribute to ASD through rare protein-truncating variation, and (2) on the autosomes, the ASD liability contributed by common variants dwarfs the combined contribution of *de novo* and rare inherited variants [50–52]. We might therefore hypothesize that the X could also harbor common, small-effect alleles, only beginning to be identified [53, 54]. If there were, for example, 150 recessive variants on the X with a frequency of *q* = 0.1, imparting an odds ratio of 1.012 with corresponding *β* = 0.00428 and *δ* = −0.00385, they would combine to shift the male mean sufficiently to account for the entirety of the observed prevalence difference (−150 × *δ* = 0.577; Figure 4B). At the same time, they would increase broad-sense heritability by values small enough (0.27% in females and 2.4% in males) that they would be difficult to detect in classical family-based heritability studies (Supplementary Methods section 1.2.3).

This example shows how common (and rare) variants on X could explain the entirety of the prevalence difference for ASD. Indeed, an analysis of male pattern baldness [55] provides a natural example of how differential sex prevalence arises. Nonetheless, this mechanism for sex differential prevalence for ASD cannot be proven or refuted without further genetic data and analyses.

## 3 Discussion

The fact that generally females have two copies of every X-linked gene whereas males have only one has created challenges for modeling genetic variation on the X for a long time. Because of differences from the autosomes, even calling genetic variation on the X can be more error-prone [56, 57]. In this manuscript, we focus on modeling rare exonic variation on the X, using data from ASD as an exemplar. We develop a new model for associating genes on the X with a binary phenotype, identify ASD-associated genes by applying it to a large exome-sequenced cohort, introduce new theory to explain genotypic effects and sex differences in the prevalence of binary phenotypes, and highlight the implications of these models for ASD.

ASD makes for an interesting case study. It has a strong male-biased sex prevalence, and prior sequencing studies have largely associated ASD with autosomal genes and been driven by *de novo* variation. Because of differences in mutation rates on the paternal and maternal gametes, we expect daughters to have far more *de novo* mutations on the X than sons, and we see this in our data (Table 2). When we apply QXL-TADA, we find 9 genes on the X associated with ASD at FDR < 0.05 and 9 more at FDR < 0.2, with PTVs providing the bulk of the ASD associations. Furthermore, there is evidence that a substantially larger number of genes could be linked with ASD in future studies, as our estimate of the proportion of non-null loci for PTVs on the X is over 18%.

At first blush, the notion that the X chromosome harbors a substantial number of genes and variants that contribute to ASD may seem inconsistent with our common understanding of “X-linked genetics,” in which one expects to find families with many affected males, few affected females, and with “transmission” of phenotype via the female line (*e.g*. hemophilia or X-linked intellectual disability). This intuition, however, derives from alleles with penetrance near 100% in males. While ~150 genes could harbor PTV variation associated with ASD, we find only 31 with an estimated penetrance greater than 50% in males. Thus, the majority of X genes cannot be considered “Mendelian” causes of ASD. Furthermore, this intuition ignores the potentially larger liability imparted by common variation on the X [50, 54].

One well-known phenomenon that our model does not include is X chromosome inactivation (XCI), which can influence the phenotypic expression and perceived inheritance patterns of X-linked disorders in heterozygous females. Typical random XCI results in a mosaic pattern with approximately 50% of cells expressing each allele. Skewed XCI patterns have been observed for two converse reasons: either the skewing contributed to ascertainment by causing sufficient symptom severity to bring the individual to clinical attention, or preferential inactivation of the X chromosome carrying the pathogenic variant ameliorated disease severity [58–60]. The possibility of different levels of XCI skew across genes on the X, the impact of ascertainment mechanisms that cause observed XCI to vary, and its impact on trait variation [61] make it challenging to build this feature into QXL-TADA.

Building on the mathematics which underlie QXL-TADA, we also develop the general theory of how X-linked variation of any additive or non-additive effect influences a phenotype and describe its implications for sex-differential prevalence. This theory has implications for the biology of autism. Various hypotheses have been proposed to explain the prevalence difference between the sexes in ASD [5–11], ranging from the liability threshold model [12, 62] to other aspects of biology, including sex hormones and sex differences in gene expression. While many influences are likely involved, our results suggest that one contributing factor could reside in genetic variation on the X, its inheritance patterns, and its effects on phenotype.

## 4 Methods

### 4.1 Cohort details

This study combines samples from the Fu et al. (2022) [21] autosomal exome study of ASD with two new cohorts. From the published study, we include four family-based trio datasets: 1) the “ASC B14” dataset, containing samples from Autism Sequencing Consortium batches 1-14 as well as the Simons Simplex Collection, 2) a smaller “ASC B15-16” dataset, containing ASC batches 15 and 16, 3) the “SPARK Pilot” exome release, and 4) the SPARK.27k.201909 exome release, called “SPARK main freeze” in Fu et al. (2022) but called “SPARK WES1 GATK” here to differentiate it from subsequent SPARK releases. We include all samples from these datasets with the exception of ASD proband 05C48707 from ASC B14, which we drop due to an aberrantly high number of *de novo* variants on the X. We do not include the 559 samples included in Fu et al. (2022) as “previously published variants,” as inherited variants are not available for these samples. We also include two case-control datasets from the published study: 1) the Danish iPYSCH cohort [22] and 2) the Swedish PAGES cohort (although this study uses a version of the PAGES data that was jointly processed with the ASC B14 dataset on GRCh38 rather than lifting over GRCh37 variants from Satterstrom et al., 2020 [22]).

The two new cohorts are 1) the “ASC B17-21” dataset, containing ASC sequencing batches 17 through 21, and 2) the “SPARK iWES v2” exome release. We did not include samples from SPARK iWES v2 that overlapped the published SPARK WES1 GATK samples. ASC B17-21 consists of complete trios while the SPARK iWES v2 consists of both complete and incomplete trios. After quality control, we PCA-matched SPARK iWES v2 ASD cases from incomplete trios with an equal number of samples from the UK Biobank (using the R library PCAmatchR on the first three genetic PCs, and matching males and females separately) to create an additional case-control cohort. In this cohort, we kept only one SPARK ASD case per family and additionally dropped one of any case pair with a kinship coefficient (from PC-Relate implemented in Hail 0.2, https://hail.is) greater than 2^−3.5^ (~0.08839).

### 4.2 Sequencing

Production of the datasets included in Fu et al. (2022) using the genome analysis toolkit (GATK [63]) was described in that manuscript. Briefly, for each of the ASC B14 and ASC B15-16 datasets, samples were processed by aligning sequence read data to the hg38 reference genome. Variants were first called individually using local realignment by HaplotypeCaller in gVCF mode and were then called jointly using GenotypeGVCFs. Variant quality score recalibration (VQSR) was run on the joint dataset to estimate variant call accuracy. SPARK Pilot bam files were downloaded from the Simons Foundation Autism Research Initiative (SFARI), realigned to the hg38 reference genome, and then processed using this pipeline. For the larger SPARK WES1 GATK release, individual gvcf files produced by GATK were downloaded from SFARI, variants were called jointly, and VQSR was run on the resulting dataset. The PAGES case-control cohort was processed as part of the ASC B14 dataset, and the iPSYCH case-control cohort was produced separately [22].

Newly analyzed datasets were produced similarly. GATK was used to produce gvcf files for the ASC B17-21 samples, and variant calling was then conducted jointly with gnomad v4.0 (https://gnomad.broadinstitute.org/news/2023-11-gnomad-v4-0) using Hail 0.2, after which samples were delivered as a subset of the larger callset. Each chromosome of the SPARK iWES v2 data was downloaded from SFARI as a vcf-format joint callset produced by GLnexus (https://github.com/dnanexus-rnd/GLnexus) using gvcf files generated by GATK. UK Biobank samples were from the 200k UKB exome cohort and were jointly called as part of gnomad v4.0.

### 4.3 Creation of working datasets

For the data analyzed in Fu et al. [21], the same working datasets were used as described in that manuscript, and ASC B17-21 and SPARK iWES v2 working datasets were produced using a similar pipeline. Briefly, Hail 0.2 was used to process each dataset individually. Low-complexity regions (using https://github.com/lh3/varcmp/blob/master/scripts/LCR-hs38.bed.gz) were dropped, and variants were assigned specific genes and consequences by annotation with the Variant Effect Predictor (VEP) [64], prioritizing coding canonical transcripts when parsing VEP output.

Several genotype filters were applied. Genotypes were dropped if they had a depth less than 10 (except for male hemizygous regions, where a minimum of 7 was allowed) or greater than 1000. Homozygous reference calls were filtered if they had a genotype quality (GQ) below 25, and heterozygous and homozygous variant calls were dropped if they had a phred-scaled likelihood of the call being homozygous reference (PL[HomRef]) below 25 or a proportion of informative reads (counting reads supporting either the reference or alternate allele for heterozygous calls, and counting only reads supporting the alternate allele for homozygous variant calls) below 90% of the depth. Heterozygous calls were further filtered if (1) the allele balance (number of reads supporting the alternate allele/depth) was below 0.25, (2) the probability of the allele balance (assuming a binomial distribution with mean 0.5) was below 10^−9^, or (3) they occurred in a male in a hemizygous region. Additionally, any calls on the Y chromosome in females were filtered.

Variant filters were applied following genotype filters. For most datasets, variants with a call rate less than a certain threshold (10% for Fu et al. data and 80% for newly analyzed data; the threshold was able to be raised due to within-dataset homogeneity of sequencing technologies used) or a Hardy-Weinberg p-value < 10^−12^ were dropped. These filters were not applied at this stage to the SPARK-UKB casecontrol cohort; instead, to limit the data to those intervals well-covered by both datasets, variants were dropped if they fell outside intervals (using the SPARK interval file union regeneron and twist.hg38.pm50.chr prefix.bed) where both datasets had at least 85% of samples with a mean depth (across sites in the VCF) of at least 20.

### 4.4 *De novo* variant calling and quality control

Starting from each working dataset, a GQ filter of 25 was applied to each genotype and raw *de novo* variants were called, followed by application of several filters to obtain a final list of variants. For autosomal *de novo* variants from the samples analyzed in Fu et al., we simply used the published variants [21]. Those variants were called using the hl.de novo() function in Hail (with variant frequencies from the non-neuro subset of gnomAD v2.1.1 GRCh38 exomes as priors, gs://gnomad-public/release/2.1.1/liftover_grch38/ht/exomes/gnomad.exomes.r2.1.1.sites.liftover_grch38.html), and the dataset-specific filters applied are described in that manuscript. As the Fu et al. manuscript does not include variants on the sex chromosomes (and as hl.de novo() is not currently well-suited for hemizygous situations), we called *de novo* variants on the X for those samples using an updated version of the Hail caller (https://github.com/ksatterstrom/ASD-Genetics/blob/main/my_de_novo_v16.py) and then applied the same filters as applied to the corresponding autosomal calls.

The same updated caller was also used to produce the list of raw *de novo* calls for the ASC B17-21 and SPARK iWES v2 datasets. Calls were dropped if they had a maximum non-neuro subpopulation frequency in gnomAD v2.1.1 above 0.1%, a within-dataset frequency above 0.1%, a proband allele balance below 0.3, a depth ratio (comparing the proband depth to the sum of parent depth, adjusted for ploidy) below 0.2, or a FisherStrand value (Phred-scaled Fisher’s exact p value of strand balance) above 25. Indels with a pAB value less than 0.01 were dropped. Dataset-specific filters were then applied.

For ASC B17-21, calls were required to be labeled “high” confidence by the caller, or to be “medium” confidence with an allele count of 1 in the dataset. Calls were additionally required to have an allele-specific quality score at least 200 (single nucleotide variants, *i.e*. SNVs) or 400 (indels) and a variant call rate at least 0.97 (SNVs) or 0.99 (indels). Non-hemizygous calls were required to have ReadPosRankSum and BaseQRankSum absolute values no greater than 2.8 (SNVs) or 1.8 (indels), as well as MQRankSum absolute values no greater than 0.6 (SNVs) or 0.2 (indels) (hemizygous calls did not have these values). In addition, calls were dropped if they had come from sites with greater than four different alleles.

For SPARK iWES v2, because of the size of the dataset, we adjusted the maximum allele count for “medium”-confidence indels in the caller to be 10 rather than 5. Calls were then required to be labeled “high” confidence by the caller, or to be “medium” confidence with an allele count no greater than 10 in the dataset. Calls were additionally required to have an allele-specific quality score at least 100, a variant call rate at least 0.9, and neither parent with a read supporting the reference allele at that site. Calls were also dropped if they came from a site with more than six different alleles in the dataset or if they occurred more than nine times (this dropped one synonymous variant and one missense variant with no MPC score).

Following these filters, for both datasets, *de novo* variants were limited to one variant per person/gene. Individuals were then dropped if they had significantly more coding *de novo* calls than their dataset mean (via Poisson test); this led to one proband and two siblings from SPARK iWES v2 not being included here.

### 4.5 Gene-level genotype details

Dataset-specific filters were applied when processing the inherited data. For all datasets, a GQ minimum of 25 was required for all genotypes, and variants with “ExcessHet” in the filters field were dropped. Variants were also required to have a population reference frequency (using the gnomAD v2.1.1 non-neuro subset for the Fu et al. datasets as in that manuscript, and the gnomAD v2.1.1 maximum non-neuro subpopulation frequency for ASC B17-21 and SPARK iWES v2) no greater than 0.1%. Variants were dropped if they had a within-dataset frequency above 0.1% (for the larger ASC B14, SPARK WES1 GATK, and SPARK WES1 GATK datasets) or above 1% (for the smaller ASC B15-16, SPARK Pilot, and ASC B17-21 datasets).

For the ASC B14 dataset, autosomal sites were dropped if they had a VQSLOD < 5.13 or an allele-specific ReadPosRankSum < −0.8 (SNVs) or −8 (indels). X non-PAR sites were filtered if they had a VQSLOD < −1.5 or an allele-specific ReadPosRankSum < −8. All calls were subject to filters requiring QD at least 3 for SNVs and at least 1 for indels, as well as allele-specific SOR no greater than 3. In addition, on the X non-PAR, calls were dropped if they came from a site with VQSLOD < 5.13 or if the variant was an SNV with allele-specific ReadPosRankSum < −0.8 *unless* the only variant in the trio came from a father who did not have a daughter in the dataset, in which case these thresholds were not applied. This adjustment was necessary to account for the fact that GATK gives a variant a higher quality score if it appears in more than one person, and any X non-PAR variant in a father with daughters in the dataset would be transmitted and therefore observed multiple times while any variant in a father without daughters would not be.

Similarly, for the SPARK GATK WES1 dataset, autosomal sites with VQSLOD < 6.01 and X non-PAR sites with VQSLOD < −0.75 were filtered. X non-PAR calls were then dropped if they came from sites with VQSLOD < 6.01 unless the only variant in the trio came from a father without a daughter in the dataset. For the other two datasets from Fu et al., requiring the site to “PASS” VQSR was the only filter necessary.

For the ASC B17-21 dataset, variants were dropped if they came from a site with more than 4 alleles or had a call rate < 0.97 (SNVs) or 0.99 (indels), depth ratio < 0.1, or VQSLOD < 3. Variant calls in probands specifically were then required to have a higher VQSLOD threshold of 4.2. For the SPARK iWES v2 dataset, variants were dropped if they had call rate < 0.99 or allele quality < 460.

Mutation rates were calculated using a sequence-context model based on gnomAD v4.1 mutation rate estimates (see https://github.com/ksatterstrom/ASD-Genetics/blob/main/Mutation_rate_calculations_for_ASC_chrX.ipynb).

### 4.6 X-linked loci increase the prevalence of ASD in males relative to females

Understanding how X-linked loci *per se* can contribute to prevalence differences begins with a detailed analysis of single-locus genetics. Consider a single locus on the X chromosome in Hardy-Weinberg equilibrium with two alleles, 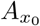 and 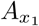, where the frequency of 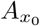 is *p* and the frequency of 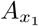 is *q* = 1 − *p* (with *p* ≥ *q*). Call *G* the genotype of an individual. Diploid females have three possible genotypes, 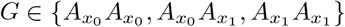, and males have two, 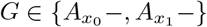.

Let *P* represent the phenotype of an individual and assume that *P* is zero-centered in diploid individuals, *i.e*. the mean of *P* is zero for females. Left superscripts are used throughout to denote quantities in males (*m*) and in females (*f*). We can then derive the following relationships [65].

Define the genetic effect of genotypes 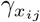 as the average phenotype of individuals with those genotypes (for a binary trait such as ASD, this is the same as the liability).

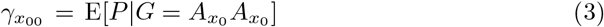

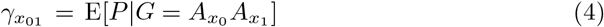

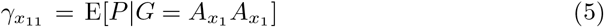

As we will show below, if the mean phenotype in females is 0, and there is any dominance, then the mean phenotype in males necessarily differs from 0. We will assume throughout that the difference in average phenotype between homozygous females is the same as the difference in average phenotype between hemizygous males. Thus, in males

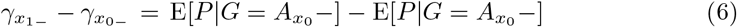

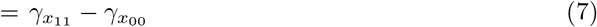

Therefore, the genetic effects of homozygotes differ between males and females by at most some constant *C*.

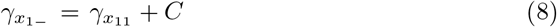

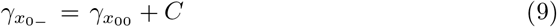

Define the allelic effect as the average phenotype of diploid individuals (females) who possess a given allele.

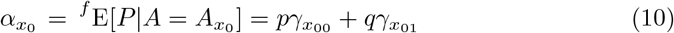

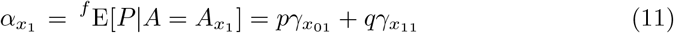

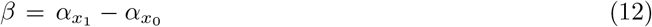

Thus, *β* is defined as the difference in allelic effects between the two alleles. Because the square of this difference (see below) is proportional to the additive variance, *β* itself is often, perhaps confusingly, also referred to as the “additive effect,” as we do in the Results.

Define the dominance deviation of a genotype 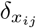 as the difference between the genetic effect of that genotype from the sum of its individual allelic effects in diploid (female) individuals. We use the facts that 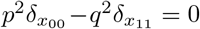 and that the expectation of the dominance deviation in females is zero (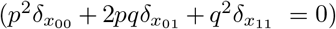) to write the dominance deviations for both homozygotes in terms of 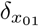.

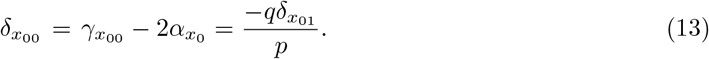

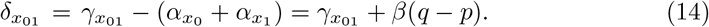

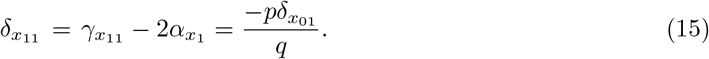

Because all three dominance deviations 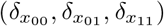 can be written in terms of a single deviation, 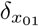, we find it convenient to do so, and denote that single dominance deviation as 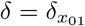 in the Results. Importantly, note that whenever there is dominance at this locus, the mean phenotype in males is not zero.

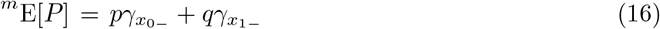

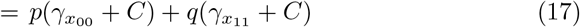

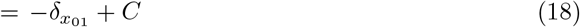

If there are some other genetic or environmental factors that interacted with this locus in a sex-specific way, a natural method to model this would be through *C*. Here we will assume that there are no interactions between this locus and any other factor, and assume *C* = 0. The additive, dominance, and genetic contributions to phenotypic variance due to this locus in females are

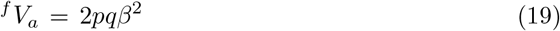

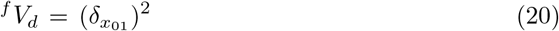

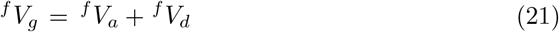

Thus, for loci on the X chromosome, the dominance contribution to variance in females is equal to the expected phenotype in males squared. For males, we find the additive contribution to variance is twice that of females

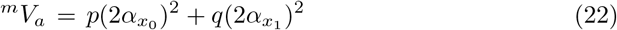

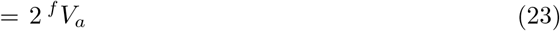

The dominance contribution in males may be larger or smaller than the female dominance. Unlike females, there is covariance between the additive and dominance contributions in males.

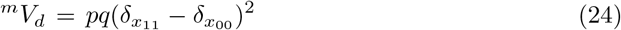

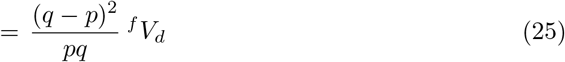

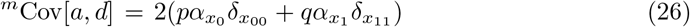

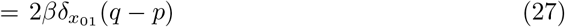

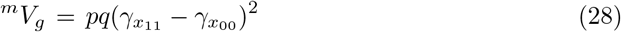

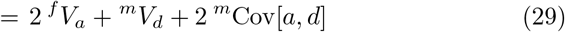

## Supplementary information

Supplementary methods and results can be found in the Supplementary Materials.

## Acknowledgments

This work was supported by NIH Grant RF1 AG071170, R01 MH129725 (K.R.), R01 MH129724 (J.D.B.), R01 MH129722 (M.J.D.), as well as prior ASC funding (U01 MH100233 and U01 MH111661). Additional support was provided by the Seaver Autism Center for Research and Treatment, the Seaver and the SWT Foundations. The funders had no role in study design, data collection and analysis, decision to publish, or preparation of the manuscript.

## Declarations

### Ethics approval and consent to participate

Not applicable.

### Consent for publication

All authors consent to publication. No previously copyrighted material included.

### Data availability

Not Applicable.

### Materials availability

Not Applicable

### Code availability

QXL-TADA code freely available from https://gitlab.com/gadhatem/david_cutler_lab_x_chromosome/

### Author contribution

All authors participated in the derivation, writing, and editing of this work.

